# Functional brain connectivity in early adolescence after hypothermia-treated neonatal hypoxic-ischemic encephalopathy

**DOI:** 10.1101/2024.05.31.24308185

**Authors:** Gustaf Håkansson, Katarina Robertsson Grossmann, Ulrika Ådén, Mats Blennow, Peter Fransson

**Affiliations:** Department of pediatrics, Karolinska University Hospital, Stockholm, Sweden; Department of Clinical Intervention and Technology, Karolinska Institutet, Stockholm, Sweden; Department of Women’s and Children’s health, Karolinska Institutet, Stockholm, Sweden; Department of Clinical Neuroscience, Karolinska Institutet, Stockholm, Sweden

## Abstract

**Aim:** Neonatal hypoxic-ischemic encephalopathy (HIE) injures the infant brain during the basic formation of the developing functional connectome. This study aimed to investigate long-term changes in the functional connectivity (FC) networks of the adolescent brain following neonatal HIE treated with therapeutic hypothermia (TH).

**Method:** This prospective, population-based cohort study included all infants (n=66) with TH-treated neonatal HIE in Stockholm during 2007-2009 and a control group (n=43) of children with normal neonatal course. Assessment with resting-state functional magnetic resonance imaging (fMRI) was performed at Karolinska Institutet, Stockholm at age 9-12 years.

**Results:** fMRI data met quality criteria for 35 children in the HIE-cohort (mean [SD] age at MRI: 11.2 [0.74] years, 46% male) and 30 children in the control group (mean [SD] age at MRI: 10.1 [0.78] years, 53% male). Clinical neurologic symptoms were present in 40% of children in the HIE-cohort. Non-parametric statistical analysis failed to detect any significant (p<0.001) alterations of FC networks in the HIE-cohort, nor between children in the HIE-cohort with or without neurological sequelae.

**Interpretation:** HT-treated HIE could not be associated with persistent alteration of the functional connectome. This indicates a notable resilience of the functional architecture of the brain against this type of early brain injury.

## Introduction

Neonatal hypoxic-ischemic encephalopathy (HIE) affects the infant brain during the time of basic formation of the developing functional connectome. The functional connectivity (FC) networks that comprise the connectome emerge as rudimentary networks already in the fetal brain (1). By full term, primary sensory areas have formed more mature networks, while higher-order networks are present in an immature state (1,2). During child development, these networks need to undergo dramatic changes in order to fully integrate to a mature functional connectome (1). Resting-state functional magnetic resonance imaging (rs fMRI) provides an opportunity to investigate networks by utilizing the slow fluctuations in the blood oxygen level dependent (BOLD) signal to calculate temporal correlations between brain regions (3).

HIE affects approximately 1-3 per 1000 live born children in countries with developed health care and is a common cause of severe neurological disability (4). The acute event evolves into a neuro-detrimental process that can persist long after the initial insult (5). HIE can also negatively impact neuroplasticity, which is essential for both normal brain development and recovery following brain injury (6). The severity of encephalopathy is often assessed using a three-grade clinical score, such as the Sarnat score (7). Treatment with therapeutic hypothermia (TH) reduces the risk for disability and has been implemented as Standard of care for moderate to severe HIE in most high resource settings (8). However, even with TH-treatment, most affected children exhibit some neurologic abnormality when assessed at school age (8,9).

A few studies have identified aberrant FC networks in infants with neonatal HIE (10–14), and one study have investigated effects at age 6-8 years (15). By early adolescence, FC network development has typically reached a stable modular organization after which no significant network reorganization is expected (1). The aim of our study was to examine the effects of TH-treated neonatal HIE on the FC networks of the brain during early adolescence.

## Method

### Participants

All term or near-term ( ≥34 weeks gestational age) infants in Stockholm from January 2007 to December 2009 who met clinical criteria for neonatal HIE and received TH-treatment were included in the cohort. Children with genetic and/or metabolic disorders known to have neurological effects, were excluded. This cohort and the clinical outcomes have been described previously by Robertsson Grossmann et al (2022) (16). From the originally 66 included children, one was excluded due to a genetic disorder and an additional 13 were ineligible for this follow-up study due to being either deceased (n=8) or moved abroad (n=5) (see Figure S1 for flowchart). The remaining 52 children were invited to participate. A control group of 43 children participated in the same follow up protocol. The controls were singleton, term-born in Stockholm Sweden with an uneventful neonatal period (5-min Apgar score > 3) that originally were recruited through random identification from the Swedish Medical Birth Registry for a separate study (17). Our study was approved by the Ethical Review Board in Stockholm, Sweden (2009/735-31/4, 2010/850-31/1, 2012/617-32, 2016/1921-32, 2019-01447, 2020-03318) and conformed to the Declaration of Helsinki. Written, informed consent was obtained from all care givers and children gave assent to participation.

### Data acquisition

Data collection was performed at Karolinska Institute and Karolinska University Hospital, Stockholm, Sweden with neonatal and medical information collected prospectively. Follow up of the HIE cohort was performed from January 2018 to December 2019 at age 10-12 years. Medical assessment was performed by an experienced pediatric neurologist. Intelligence was assessed using Wechsler Intelligence Scale for Children, fifth edition (18) (WISC-V, Swedish version), performed by either an experienced child psychologist or a psychologist under supervision. Clinical diagnoses were retrieved from the participants’ electronic medical records.

MR images of the brain were acquired using a Sigma 3T MR Scanner (GE Healthcare) at the MR-Centrum, Karolinska Institute. No sedation was used. Participants were prepared for the scanning with an optional information movie and familiarization using a mock scanner. At least one parent was present with the child before scanning and observing from the monitor room. During anatomical MRI acquisition all participants were offered to watch a movie or listen to music of their choice. For the resting state fMRI session, the participants were instructed to remain still and keeping their eyes open. T1-weighted two-dimensional spin-echo images (TE = 2,7 ms, flip angle 12 degrees, slice thickness 1.0 mm) were obtained in three planes with a 64-channel head coil. For the resting state fMRI session, 300 T2*-weighted whole-brain echo-planar image (EPI) volumes were acquired over 10 minutes (TR = 2.0 s, TE = 30 ms, flip angle 70 degrees, 3x3x3 mm voxel size).

### Image Preprocessing and Denoising

Both functional and anatomical images were minimally preprocessed using fMRIPrep (19) 20.2.4. Anatomical T1-weighted images were corrected for intensity non-uniform signal intensity with N4BiasFieldCorrection (20). After skull-stripping with antsBrainExtraction, the T1-weighted references were normalized to a pediatric template (7.5 to 13.5y age range [TemplateFlow ID: MNIPediatricAsym:cohort-4] (21)) using volume-based spatial normalization through nonlinear registration with antsRegistration (22) 2.3.3.

A visual quality control was performed and image quality metrics for resting-state fMRI data were extracted using MRIQC (23) 0.16.1. For most subjects, motion artifacts were pronounced at the end of the rs-fMRI sequence. We therefore decided to exclude the last minute of the data recorded, resulting in a 9-minute BOLD sequence (270 volumes) for all subjects. Criteria for data exclusion related to excessive subject head motion was set to frame-wise displacement (FD) > 0,5 mm on a maximum of 20% of image volumes or DVARS > 1.5 SD.

In fMRIPrep, a functional reference volume and its skull-stripped version was generated using a median of a motion corrected subset of volumes. The functional reference volume was co-registered to the T1-weighted image volume with FLIRT (24) (FSL 5.0.9) using a boundary-based registration cost-function (25). The co-registration was performed using nine degrees of freedom to account for remaining spatial distortions in the functional reference volume. Functional images were resampled to native space using a fixed-body model (three translational and thee rotational movement regressors) that was estimated from the reference volume using MCFLIRT (26) (FSL 5.0.9). After slice-time correction to 0.976s using 3dTshift from AFNI, the resampled functional images were further resampled into a pediatric standard space (MNIPediatricAsym:cohort-4) (21).

Signal denoising of the BOLD signal time-series with respect to non-neuronal signal sources was performed using principal component analysis-based noise regression. For this, we used CompCor (27) to generate three probabilistic masks (cerebrospinal fluid (CSF), white matter (WM) and combined CSF+WM) in native anatomical space. This was done using principal component analysis such that the retained components’ time series are sufficient to explain 50% of variance across the mask. Preprocessed BOLD time-series in MNI space were spatially smoothed with an isotropic, Gaussian kernel of 6 mm FWHM (full-width half-maximum). The denoising regression was performed in one single regression model to avoid reintroduction of artifacts (28) with the following variables: six rigid body realignment parameters, top five anatomical CompCor decompositions, motion outliers (frames exceeding 0.5 mm FD), FD and cosine filter (128s cut-of). The data was low pass filtered at 0.1 Hz, no high pass filter was used aside from the cosine filter.

Group level spatial brain FC maps (with the HIE cohort and control group treated as a single dataset) were calculated in Nilearn using canonical ICA (29) with 20 fixed components. ICA was chosen rather than a static, parcellation-based delineation of the brain to increase methodological flexibility for any cortical reorganization and/or recruitment of cortical areas outside the standard location. Canonical ICA improves the ICA-analysis by adding a canonical correlation analysis to the ICA-based pattern extraction for identification of a common data subspace and thresholding the independent components based on the absolute value of voxel intensity (29).

### Statistical analysis

Comparison of FC between children with HIE and controls was performed using dual regression where the spatial maps derived from the canonical ICA were used to generate subject-specific versions of both spatial maps and associated timeseries in a two-step regression analysis (30). Testing for group-differences was performed using non-parametric testing with 5000 permutations with correction for multiple comparisons using the threshold-free cluster enhanced (TFCE) technique (31,32).

As noted in Table 1, age at MRI differed significantly between the groups. To address a possible introduction of bias, we therefore performed an additional dual regression analysis in which we included a covariate pertaining to group differences for age at scan in the dual regression model. Similar to the previous model, significance of group differences was performed using non-parametric testing (5000 permutations, multiple comparison correction using TFCE). Results were thresholded for a minimal cluster size of 10 voxels and p-values of all significant results were further corrected for multiple comparison of all FC components using Bonferroni correction.

**Table 1.** Characteristics of study population stratified by exposure to hypothermia-treated neonatal hypoxic-ischemic encephalopathy.

| Characteristic | HIE (n=35) | Control (n=30) | P value |
| --- | --- | --- | --- |
| Female, No (%) | 19 (54.3) | 14 (46.7) | 0.55 |
| Male, No (%) | 16 (45.7) | 16 (53.3) |  |
| Gestational age at birth,<br>median (IQR), wk | 40.6 (39.2 – 41.2) | 40.1 (39.4 – 41.1) | 0.80 |
| Birth weight, median (IQR), g | 3500 (3326 - 3984) | 3590 (3329 - 3738) | 0.88 |
| Age at WISC-V, median (IQR), y | 11.3 (10.5 – 12.8) | 12.1 (11.9 – 12.5) | < .001 |
| Age at MRI, median (IQR), y | 11.4 (10.5 – 11.8) | 10.0 (9.6 – 10.6) | < .001 |
| Frame-wise Displacement<br>during MRI, mean (SD) mm | 0.19 (0.13) | 0.15 (0.08) | 0.14 |
| DVARs (standardized)<br>during MRI, mean (SD) mm | 1.14 (0.068) | 1.16 (0.097) | 0.32 |

Abbreviations: IQR, interquartile range; SD, Standard Deviation; WISC-V, Wechsler Intelligence Scale for Children version 5; MRI, Magnetic Resonance Imaging.

A subgroup analysis was performed in which we divided the HIE-cohort into two groups based on clinical outcome to examine if children with neurologic sequelae hade more alterations than normal development, or if evidence for network reorganization affecting the functional outcome could be detected in the group of HIE exposed children with normal development. Subjects fulfilling any of the following criteria were classified as “unfavorable” outcome: IQ < 85, Cerebral Palsy (CP), epilepsy, Central Visual Impairment (CVI), cerebral hearing impairment, Autism Spectrum Disorder (ASD), Attention Deficit Hyperactivity Disorder (ADHD) and Developmental Coordination Disorder (DCD). Remaining children with HIE were subsequently classified as “favorable” outcome. Testing for subgroup-differences was done on the two subgroups with non-parametric testing similar to the main group analysis.

## Results

### Study sample

From the eligible HIE-cohort (n=52), four children declined all participation in the study for personal reasons. An additional eight children specifically declined assessment with MRI, five of these due to worry about being in the scanner, two due to severe CP and one due to not having time. In the control group one participant did not complete the whole MRI protocol and two participants were excluded due to the detection of cerebral abnormalities (one cerebellar arachnoid cyst and one cortical cavernoma). An additional 15 participants were excluded based on set criteria for excessive head motion during rs-fMRI, five from the HIE-cohort and ten from the control group (see Figure S1 for an overview flowchart and Table S3 for sensitivity analysis). Characteristics of the final study sample (n=65), consisting of 35 children in the HIE-cohort and 30 children in the control group, is presented in Table 1. In the HIE-cohort, clinical neurologic diagnoses before the follow-up were: 8 children (23%) with neurodevelopmental diagnose (i.e. ADHD, ASD and/or DCD), 3 children (9%) with CP, 1 child (3%) with neurologic hearing impairment and 1 child (3%) with CVI (Table S2). HIE grading was moderate (grade II) in 33 (94%) and severe (grade III) in 2 (6%) of the exposed children (Table S3).

### Group analysis of functional connectivity

20 fixed independent components were extracted from the combined bold dataset, of which five were categorized as representing noise or artifacts. The remaining 15 FC networks were in general agreement with known brain networks from previous literature (33) hierarchically representing nine well-established networks: Occipital (visual) Network, Cerebellar Network,

Hippocampal/Brainstem Network, Pericentral (somatomotor) Network, Pericentral (auditory) Network, Lateral Frontoparietal (control) Network, Dorsal Frontoparietal (attention) Network, Medial Frontoparietal (Default Mode) Network, Midcingulo-Insular (salience) Network (Figure S2).

The non-parametric permutation tests revealed a total of 25 significant clusters residing in gray matter (p<0.05, uncorrected for multiple statistical tests) in four of the 15 FC networks with a majority residing in the cerebellar network (Table S1, Figure S3A). After subsequent additional correction for multiple comparisons across individual networks (15 two-sided tests, corrected significance threshold: p<0.00167), the medial visual network showed a decrease of FC in the right dorsolateral prefrontal cortex (DLPFC) for the HIE cohort compared to the control group (Figure S3B). Excluding the participant with CVI from analysis did not alter this result. However, including age at scan as a co-variate of no-interest in our dual regression model resulted in this finding no longer being significant. From the 25 significant clusters in the original analysis (before correction for multiple comparisons), only 11 findings in two functional networks were significant after inclusion of age as a covariate. Using AtlasReader (34), all clusters were anatomically labeled by three different atlases (Automated Anatomical Labelling (AAL), Harvard-Oxford and Desikan Kiliany). Combined with visual inspection, we deemed four of these clusters to be mainly localized in cerebral white matter. The remaining 7 cluster differences consisted of decreased FC in the HIE cohort in the left lateral Frontoparietal Network and the Cerebellar Network (Table 2, Figure 1). None of these remained significant after correction for multiple comparisons.

**Fig.1.**
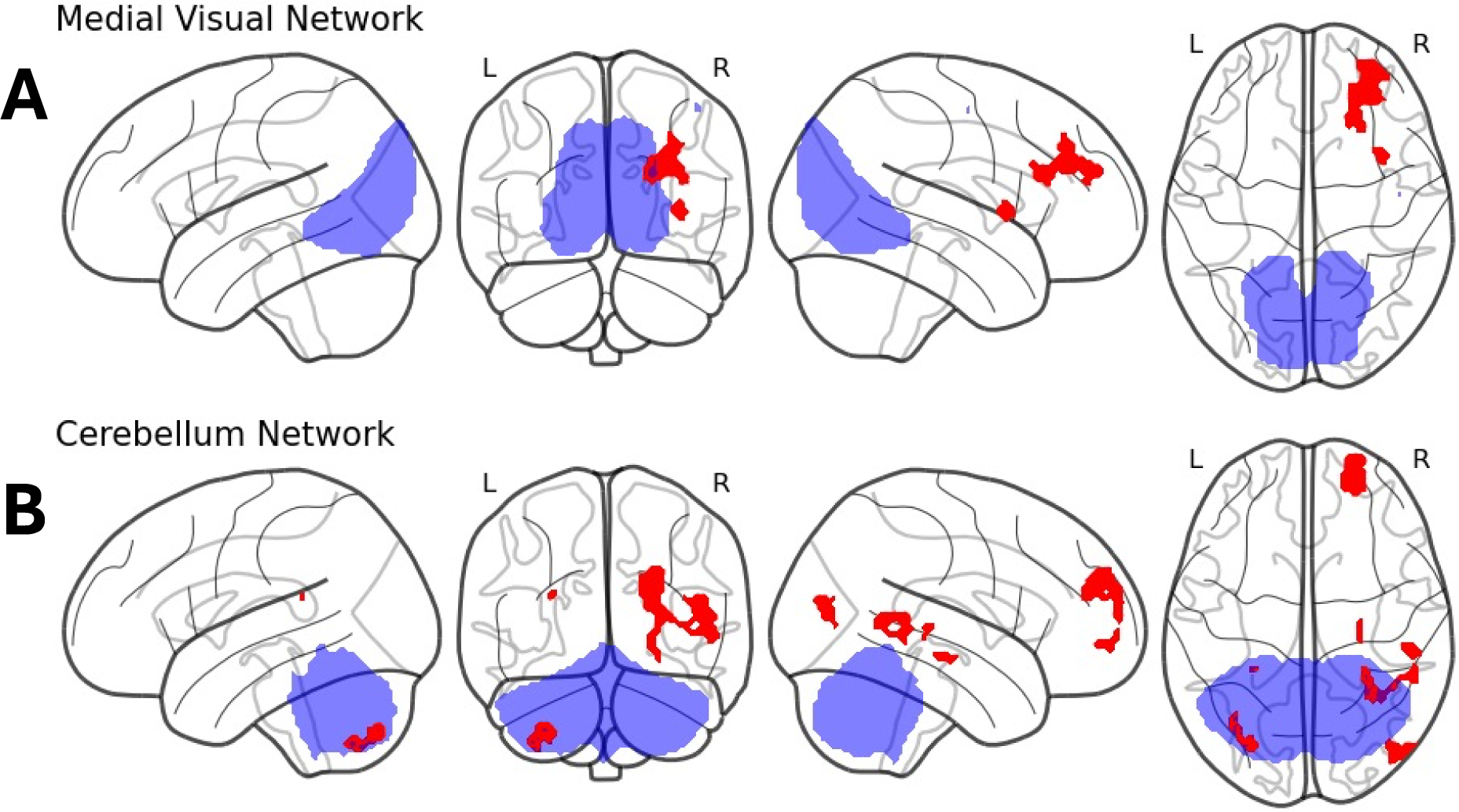
Differences in functional connectivity between groups with and without exposure for hypothermia-treated neonatal hypoxic-ischemic encephalopathy corrected for age difference. Results from non-parametric testing (p<0.05, age corrected, voxel-wise threshold free cluster enhancement corrected (TFCE), minimal cluster size = 10) on whole brain dual regression of the 15 ICA-derived networks (see Figure S2) detected clusters of decreased FC in children with hypothermia-treated HIE (N=35) compared to the control group (N=30) in the left Lateral Frontoparietal network (A) and the Cerebellar network (B). All significant changes (before correction for multiple comparisons) are colored red with seed networks are colored blue (transparent).

**Table 2.** Age corrected functional connectivity differences between groups with and without exposure for hypothermia-treated neonatal hypoxic-ischemic encephalopathy.

| Network | Location | Hemisphere | Volume<sup>a</sup><br>(mm<sup>3</sup>) | P value<sup>b</sup> | Peak<br>x<sup>c</sup> | Peak<br>y<sup>c</sup> | Peak<br>z<sup>c</sup> |
| --- | --- | --- | --- | --- | --- | --- | --- |
| Left lateral FPN | Superior Temporal | Right | 2684 | 0.01 | 51.5 | -36.5 | 11.5 |
| Left lateral FPN | Superior Temporal | Right | 8 | 0.0498 | 59.5 | -22.5 | 11.5 |
| CN | Frontal Pole | Right | 3144 | 0.013 | 23.5 | 59.5 | 27.5 |
| CN | Cerebellum | Left | 872 | 0.0224 | -30.5 | -78.5 | -48.5 |
| CN | Superior Temporal | Right | 712 | 0.0434 | 49.5 | -26.5 | 5.5 |
| CN | Hippocampus | Right | 232 | 0.0432 | 27.5 | -22.5 | -10.5 |
| CN | Middle Temporal | Right | 16 | 0.0492 | 39.5 | -54.5 | 19.5 |

<sup>a</sup>Thresholded for minimal cluster size of 10 voxels
<sup>b</sup>TFCE-corrected
<sup>c</sup>MNI coordinates
Abbreviations: FPN, Frontoparietal Network; CN, Cerebellar Network; VN, Visual Network; TFCE, Threshold Free
Cluster Enhancement; MNI, Montreal Neurological Institute and Hospital.

### Subgroup analysis

All 35 children in the HIE-cohort and a total of 22 children in the control group completed cognitive assessment with WISC-V. Mean full scale IQ was 100 (SD 16,9) in the HIE-cohort and 112 (SD 12,5) in the control group. In the HIE-cohort, 14 (40%) of the children met the set criteria for unfavorable outcome and the remaining 21 (60%) had favorable outcome (Table S2). The two groups differed regarding gestational age at birth, with the Unfavorable outcome-group being born significantly earlier (Table 3). Non-parametric testing for differences between the favorable and unfavorable subgroups (TFCE-corrected) detected increased FC for the Favorable outcome-group between the right lateral frontoparietal network and the cerebellum bilaterally and between the dorsal frontoparietal network and the bilateral anterior cingulate and postcentral cortex (Table 4, Figure 2). None of these findings did however survive additional correction for multiple comparisons (threshold p<0.00167).

**Fig.2.**
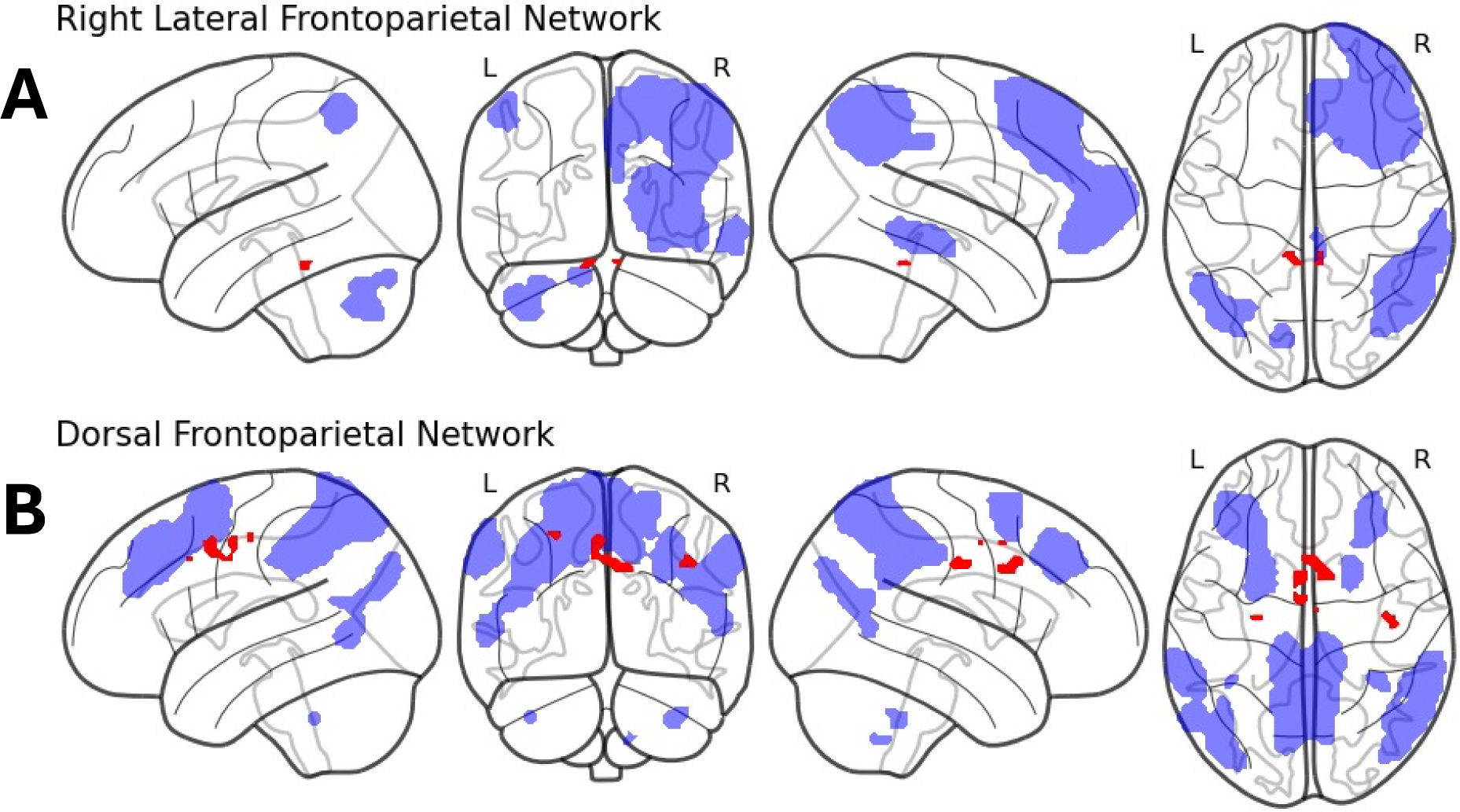
Differences in functional connectivity between children exposed to hypothermia-treated neonatal hypoxic-ischemic encephalopathy with and without neurologic sequelae. Results from non-parametric test (p<0.05, TFCE, minimal cluster size = 10) of a dual regression analysis showing clusters of increased FC in the favorable subgroup compared to the unfavorable subgroup between the right Lateral Frontoparietal Network to the cerebellum bilaterally (A) and increased FC in the unfavorable subgroup compared to the favorable subgroup between the Dorsal Frontoparietal Network to bilateral anterior cingulate and postcentral cortex (B). All significant changes are colored red, seed networks are colored blue.

**Table 3.**
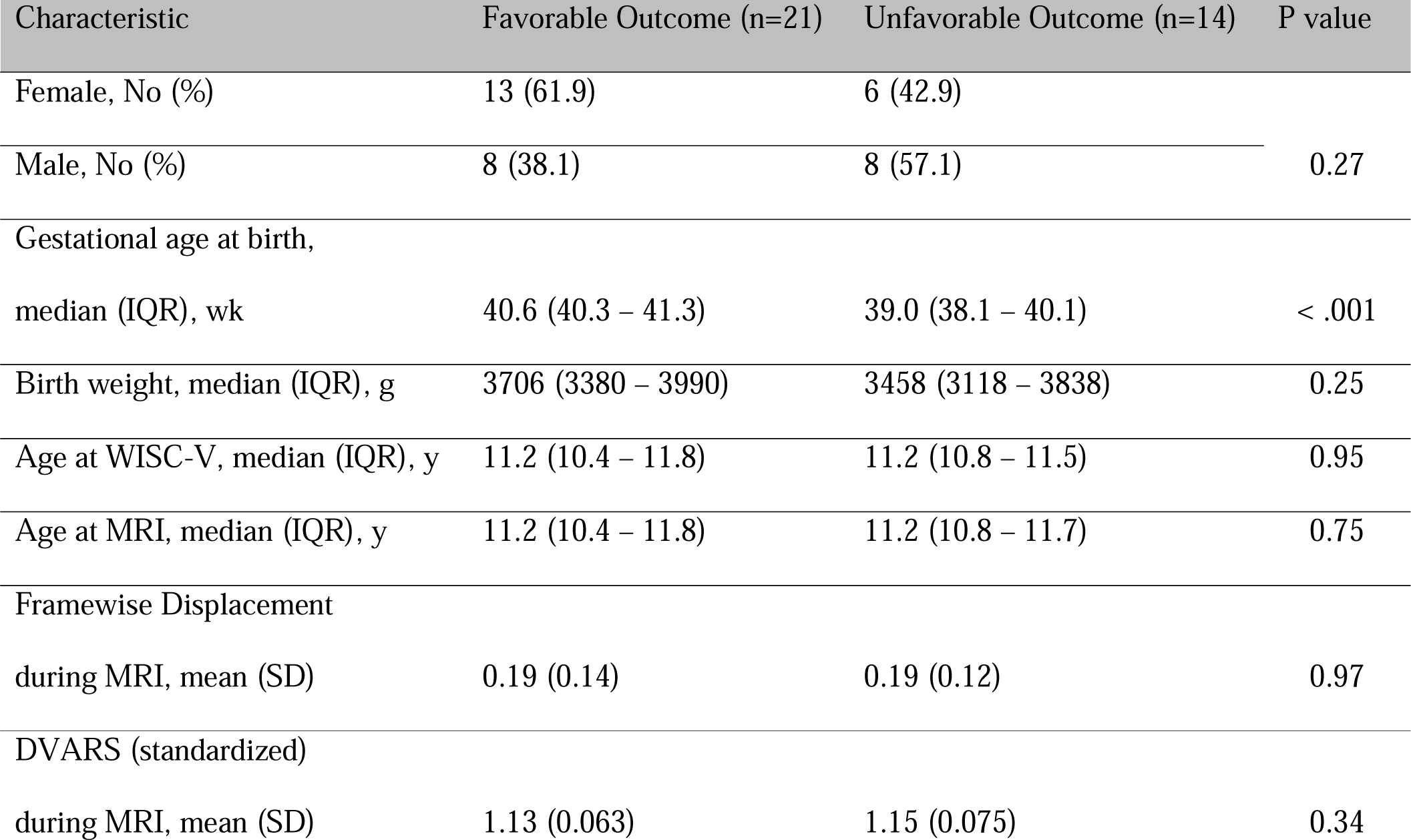
Characteristics of the HIE-cohort stratified by “favorable” or “unfavorable” outcome as determined by received clinical neurologic and/or neurodevelopmental diagnosis and/or IQ < 1,5 SD.

| Characteristic | Favorable Outcome (n=21) | Unfavorable Outcome (n=14) | P value |
| --- | --- | --- | --- |
| Female, No (%) | 13 (61.9) | 6 (42.9) | 0.27 |
| Male, No (%) | 8 (38.1) | 8 (57.1) |  |
| Gestational age at birth,<br>median (IQR), wk | 40.6 (40.3 – 41.3) | 39.0 (38.1 – 40.1) | < .001 |
| Birth weight, median (IQR), g | 3706 (3380 – 3990) | 3458 (3118 – 3838) | 0.25 |
| Age at WISC-V, median (IQR), y | 11.2 (10.4 – 11.8) | 11.2 (10.8 – 11.5) | 0.95 |
| Age at MRI, median (IQR), y | 11.2 (10.4 – 11.8) | 11.2 (10.8 – 11.7) | 0.75 |
| Framewise Displacement<br>during MRI, mean (SD) | 0.19 (0.14) | 0.19 (0.12) | 0.97 |
| DVARs (standardized)<br>during MRI, mean (SD) | 1.13 (0.063) | 1.15 (0.075) | 0.34 |

**Table 4.**
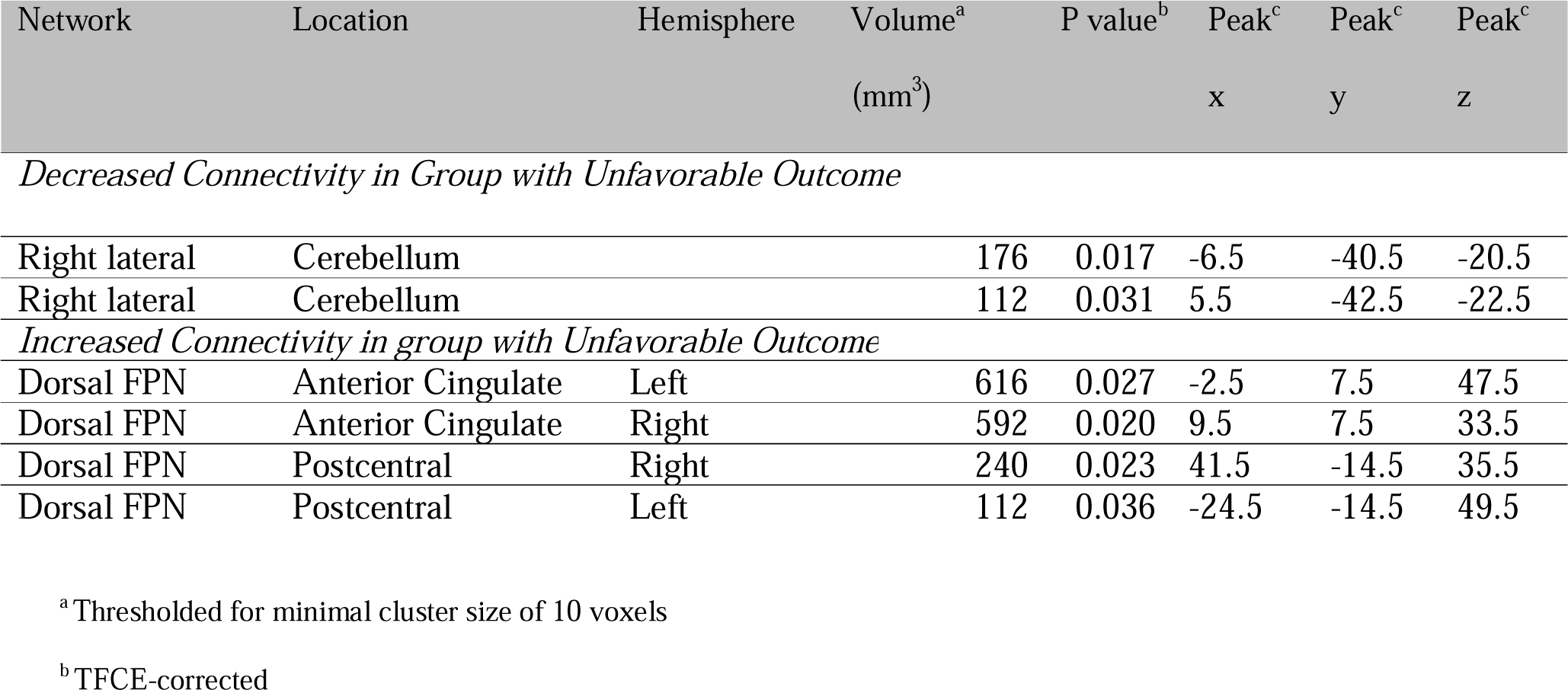
Differences in functional connectivity between children exposed to hypothermia-treated neonatal hypoxic-ischemic encephalopathy with and without neurologic sequelae.

*Decreased Connectivity in Group with Unfavorable Outcome*
|  |  |  |  |  |  |  |  |
| --- | --- | --- | --- | --- | --- | --- | --- |
| Right lateral | Cerebellum |  | 176 | 0.017 | -6.5 | -40.5 | -20.5 |
| Right lateral | Cerebellum |  | 112 | 0.031 | 5.5 | -42.5 | -22.5 |
| *Increased Connectivity in group with Unfavorable Outcome* |  |  |  |  |  |  |  |
| Dorsal FPN | Anterior Cingulate | Left | 616 | 0.027 | -2.5 | 7.5 | 47.5 |
| Dorsal FPN | Anterior Cingulate | Right | 592 | 0.020 | 9.5 | 7.5 | 33.5 |
| Dorsal FPN | Postcentral | Right | 240 | 0.023 | 41.5 | -14.5 | 35.5 |
| Dorsal FPN | Postcentral | Left | 112 | 0.036 | -24.5 | -14.5 | 49.5 |

<sup>a</sup> Thresholded for minimal cluster size of 10 voxels
<sup>b</sup> TFCE-corrected

## Discussion

In this study, we examined the resting-state FC networks in early adolescent children exposed to HT-treated neonatal HIE. In our first model (i.e. without including scan at age as a co-variate in the model), no alterations were found in a majority of the FC networks. But in four of the networks, decreased FC in the HIE-cohort were observed and most prominently in the cerebellar network to several areas located across the cortex. Early injury of the cerebellum has been linked to impaired neurocognitive development (35). It has recently been recognized as underestimated and resulting in volume reductions after neonatal HIE (36,37). However, after correction for multiple comparisons across all resting-state networks, only one of these findings remained significant, showing decreased FC in the HIE-cohort between the medial visual network and a cluster localized to the right DLPFC which is a hub in the Lateral Frontoparietal (Control) Network but also partly involved in the Dorsal Frontoparietal (Attention) Network (33).

Importantly, when including age as a co-variate of no-interest in the dual regression model, no changes were found. This null result is in line with the only other study of long term FC effects after neonatal HIE (Spencer et al, 2022, preprint) who compared FC in 22 children exposed to HT-treated neonatal HIE at age 6-8 with healthy controls (N=20) (15). Taken together, the absence of significant major alterations in FC after HT-treated neonatal HIE suggests an overall high resilience of the developing human connectome against this condition. Interestingly though, Spencer et al reports case-control differences in FC between the Attention/Cognitive Control Network and the Visual Network being significant before correcting for multiple comparisons. The common minimal findings of altered FC between the Visual Network to Networks important for attention and cognitive control indicates a possible common group effect that might be concealed due to underpower of the two studies. The dorsal stream of the visual system integrates cognitive brain functions like working memory and attention to the visual function (42,43). Specific vulnerability of the dorsal stream has been described after conditions such as perinatal brain injury (44) and can give rise to a cognitive subtype of cerebral visual impairment (CVI) with disturbed visual perception and integration that is especially common after HIE (45). This is also highlighted by findings of a reduction in visuo-spatial processing ability as well as impaired structural connectivity in TH-treated HIE (46,47). A possible connection between this condition and exposure to neonatal HIE should be further investigated by combining fMRI with testing of visual function.

Earlier studies have reported aberrant FC in the early phase of neonatal HIE. Tusor et al (2014) compared the FC (acquired within five weeks form birth) of four pre-selected FC networks in infants (N=15) with neonatal HIE and healthy controls (N=15) (11). Jiang et al (2022) specifically selected motor FC networks for comparison between infants with TH-treated neonatal HIE (N=16) and neurologically intact controls (N=11) within 11 days from birth (10). Both studies reported significant disruptions in multiple FC networks. Li et al compared atlas-based acquired network properties of infants with mild (N=12) and severe (N=12) neonatal HIE and found reduced local efficiency and clustering coefficient in the severe group within the first month of life. Finally, in two retrospective studies Boerwinkle et al (2022, 2024) correlated FC from the acute phase with clinical data at 6 months (12) and up to 42 months (mean 28.2 months) (13) in children with acute neonatal brain injury including 24 children with neonatal HIE (of which 22 were exposed to moderate to severe HIE according to Sarnat criteria). In comparison of typical versus atypical FC in four selected networks they found several associations to assessment of general development, cognition, motor function as well as mortality and epilepsy. The results from these earlier studies should however be interpreted with caution mainly due to the small sample sizes employed but taken together with our results we suggest that many of the FC changes seen in the acute or subacute stage are either reversible or recovered in later development. After early focal injury, such as stroke in the neonatal period, remarkable cortical reorganization of functions such as motor and speech has been observed using fMRI (48). Our sub-group analysis compared the FC of participants with or without neurologic sequelae but failed to identify any further significant changes or alterations suggestive of functional cortical reorganization in any of the networks.

The heterogenous impact on brain from neonatal HIE, as reflected by the broad spectrum of neurological outcome of this condition, is another possible explanation for the absence of significant findings in our study. The cortical injury pattern of neonatal HIE in the acute phase (if not total) is commonly divided into either mainly Central/Basal Ganglia - Thalamus or Watershed (border zone) injury, which has been shown to provide some prognostic information of neurologic outcome (49). These patterns arise from specific brain regions (i.e., the basal ganglia, hippocampus and primary motor cortex) being more susceptible to injury as a consequence of high metabolic activity or containment of glutamate and/or from localization to the boundaries between blood supply (e.g., border zone areas in middle-frontal and parietal-occipital cortex), depending on the course of HIE (50). The patterns of injury are however commonly overlapping, whilst at the same time many exposed children present a normal clinical MRI of the brain (51). Furthermore, the secondary (and tertiary) phases of HIE involving inflammatory processes are not detectable on MRI. It is therefore possible that individual injury patterns, along with insufficient effect size, led to a dilution of the effects on a group level. In future studies, correlating the brain FC with performance from clinical testing and increasing the study population could provide more information on function specific network alterations in these children.

### Limitations

There are several limitations to this study. First, although the current work is the largest study yet of FC in this rare condition, the sample size still limits its statistical power, especially for the subgroup analysis. We also note a significant drop-out rate from the original cohort of 66 children. The final analysis only included 40% of the original cohort of surviving children with HIE grade III and 69% of the surviving children with HIE grade II (Table S3). Furthermore, some of the children were unable to participate in the MRI acquisition only due to symptom severity (i.e. cerebral palsy) and so the cohort of children included in the final analysis is not representing the full clinical spectrum of HIE. Our results should therefore only be generalized to children with TH-treated neonatal HIE without severe CP and mainly to neonatal HIE grade II.

Second, head motion during fMRI scanning is a substantial source for noise especially at younger age (52). Exposure to neonatal HIE has been associated with unsuccessful MRI in early school age (53) and diagnoses such as ADHD, autism spectrum disorder (ASD) and epilepsy also tend to be associated with more movement (54). The control group in this study was scanned at a significantly younger age (Table 1). The mean difference in age between groups was comparatively small (but significant) and effected the results when included as a co-variate of no-interest in our dual regression model. We have therefore chosen to only present the results with age included as covariate in the main article, but due to the similarity in results with respect to earlier investigations of this condition, we include no-age corrected results as supporting material.

## Data Availability

All data produced in the present study are available upon reasonable request to the authors

## Acknowledgements

We thank the children and their parents for participating.

This study was financially supported by Stiftelsen Promobilia (F20501), Stiftlesen Sunnerdahls Handikappfond (F18/20), Drottning Silvias Jubileumsfond, Lilla Barnets Fond, Jerringfonden, Stiftelsen Kempe-Carlgrenska Fonden, HKH Kronprinsessan Lovisas Förening för Barnasjukvård, Norrbacka-Eugeniastiftelsen (866/20), Neurofonden (F2020-0009) and grants provided by Region Stockholm (ALF project and Centre for innovative medicine).

## Abbreviations

ADHD: Attention Deficit Hyperactivity Disorder
ASD: Autism Spectrum Disorder
BOLD: Blood Oxygen Level Dependent
CP: Cerebral Palsy
DCD: Developmental Coordination Disorder
FD: Framewise Displacement
FC: Functional Connectivity
fMRI: functional Magnetic Resonance Imaging
FWHM: full-width half-maximum
HIE: Hypoxic-Ischemic Encephalopathy
IQ: Intelligence Quote
MRI: Magnetic Resonance Imaging
TFCE: Threshold Free Cluster Enhancement
WISC: Wechsler Intelligence Scale for Children

