## Supporting Information for "Functional brain connectivity in early adolescence after hypothermia-treated neonatal hypoxic-ischemic encephalopathy"

**Supplemental Table 1.** Functional connectivity differences between groups with and without exposure for hypothermia-treated neonatal hypoxic-ischemic encephalopathy from data analysis with whole brain dual regression without covariates in the model.

| Network | Location | Hemisphere | Volume^a^ (mm^3^) | P value^b^ | | Peak^c^ x | Peak^c^ y | Peak^c^ z |
| --- | --- | --- | --- | --- | --- | --- | --- | --- |
| Superior SMN | Thalamus | Right | 144 | 0.032 | 3.5 | | -0.5 | 7.5 |
| Left lateral FPN | Postcentral | Left | 1416 | 0.012 | -36.5 | | -36.5 | 59.5 |
| Left lateral FPN | Inferior Temporal | Left | 1080 | 0.0098 | -48.5 | | -48.5 | -10.5 |
| Left lateral FPN | Cingulate | Left | 680 | 0.025 | -8.5 | | 33.5 | 7.5 |
| CN | Cerebellum | Left | 8656 | 0.0036 | -30.5 | | -78.5 | -48.5 |
| CN | Frontal Pole | Right | 4696 | 0.0072 | 21.5 | | 53.5 | 25.5 |
| CN | Frontal Pole | Left | 3472 | 0.023 | -26.5 | | 45.5 | 41.5 |
| CN | Lateral Occipital | Right | 1704 | 0.028 | 33.5 | | -74.5 | 21.5 |
| CN | Parietal Operculum | Right | 1456 | 0.031 | 39.5 | | -26.5 | 21.5 |
| CN | Superior Frontal | Right | 1032 | 0.031 | 15.5 | | 37.5 | 17.5 |
| CN | Supplemental Motor Area | Left | 1016 | 0.037 | -16.5 | | 11.5 | 51.5 |
| CN | Superior Temporal | Right | 864 | 0.032 | 47.5 | | -26.5 | 5.5 |
| CN | Postcentral | Left | 768 | 0.034 | -52.5 | | -20.5 | 45.5 |
| CN | Precentral | Right | 536 | 0.011 | 1.5 | | -16.5 | 77.5 |
| CN | Middle Frontal | Left | 472 | 0.040 | -30.5 | | -4.5 | 49.5 |
| CN | Cerebellum | Right | 456 | 0.022 | 5.5 | | -46.5 | -48.5 |
| CN | Lingual Gyrus | Right | 360 | 0.033 | 9.5 | | -70.5 | -8.5 |
| CN | Frontal Pole | Left | 352 | 0.038 | -22.5 | | 59.5 | -4.5 |
| CN | Cerebellum | Left | 328 | 0.030 | -8.5 | | -54.5 | -44.5 |
| CN | Precentral | Right | 240 | 0.040 | 51.5 | | -0.5 | 49.5 |
| CN | Middle Frontal | Right | 168 | 0.042 | 41.5 | | 3.5 | 45.5 |
| CN | Pars triangularis | Left | 160 | 0.040 | -40.5 | | 33.5 | 9.5 |
| CN | Middle Frontal | Left | 112 | 0.039 | -48.5 | | 23.5 | 39.5 |
| Medial VN | Middle Frontal | Right | 3712 | <0.001 | 27.5 | | 39.5 | 29.5 |
| Medial VN | Insular | Right | 400 | 0,0068 | 37.5 | | 9.5 | 3.5 |

^a^ Thresholded for minimal cluster size of 10 voxels

^b^ TFCE-corrected

^c^ MNI coordinates

Abbreviations: SMN, Sensory-Motor Network; FPN, Frontoparietal Network; CN, Cerebellar Network; VN, Visual Network; TFCE, Threshold Free Cluster Enhancement; MNI, Montreal Neurological Institute and Hospital.

**Supplemental Table 2** Clinical and cognitive outcome stratified by exposure to HT-treated neonatal HIE.

Abbreviations: WISC-V, Wechsler Intelligence Scale for Children fifth version; IQR, Interquartile Range; FSIQ, Full Scale Intelligence Quote; CP, Cerebral Palsy; ADHD, Attention Deficit Hyperactivity Disorder; ASD, Autism Spectrum Disorder; DCD; CVI, Central Visual Impairment.

| Outcome Measure | HIE (N=35) | Control (N=30) | P value |
| --- | --- | --- | --- |
| *Cognitive Outcome* |  |  |  |
| WISC-V, full scale composite score, mean (SD) | 100 (16.9) | 112 (12.5) | 0.05 |
| *Clinical Outcome* |  |  |  |
| Any neurological/neurodevelopmental diagnose and/or FSIQ < 85, No (%) | 14 (40) | 0 (0) | NA |
| Cerebral Palsy, No (%) | 3 (8.6) | 0 (0) | NA |
| FSIQ < 1,5 SD, No (%) | 6 (17.1) | 0 (0) | NA |
| ADHD, No (%) | 4 (11.4) | 0 (0) | NA |
| ASD, No (%) | 2 (5.7) | 0 (0) | NA |
| DCD, No (%) | 4 (11.4) | 0 (0) | NA |
| Hearing impariment, No (%) | 1 (2.9) | 0 (0) | NA |
| CVI, No (%) | 1 (2.9) | 0 (0) | NA |

**Supplemental Table 3** Sensitivity analysis for surviving children without genetic syndrome exposed to hypothermia-treated neonatal hypoxic-ischemic encephalopathy (n=57) stratified by included or excluded from functional brain connectivity analysis.

Abbreviations: HIE, Hypoxic-Ischemic Encephalopathy; GA, Gestational Age; BW, Birth Weight; IQR, Intra Quartile Range; WISC, Wechsler Intelligence Scale in Children; IQ, Intelligence Quote.

| Charectaristic | HIE included in analysis (n=35) | HIE excluded from analysis (n=22) | P value |
| --- | --- | --- | --- |
| GA, median (IQR), wk | 40.6 (39.2 - 41.2) | 40.5 (39.1 - 41.4) | 0.75 |
| BW, median (IQR), g | 3500 (3326 - 3984) | 3594 (3143 - 4340) | 0.52 |
| Apgar score at 10 minutes, median (IQR) | 4 (2.5 - 6) | 5 (4 - 6) | 0.34 |
| *Sarnat grade of HIE* |  |  |  |
| Grade I, No (%) | 0 (0) | 4 (18.2) | 0.015 |
| Grade II, No (%) | 33 (94.3) | 15 (68.2) |  |
| Grade III, No (%) | 2 (5.7) | 3 (13.6) |  |
| *Maximum respiratory support* |  |  |  |
| None, No (%) | 8 (25.7) | 4 (18.2) | 0.47 |
| CPAP, No (%) | 3 (8.8) | 4 (18.2) |  |
| IPPV, No (%) | 16 (47.1) | 12 (54.5) |  |
| HFO, No (%) | 7 (20.6) | 2 (9.1) |  |
| Missing, No (%) | 1 (2.9) | 0 (0) |  |
| *Worst EEG background activity:* |  |  |  |
| Continuous, No (%) | 8 (22.9) | 1 (18.2) | 0.067 |
| Discontinuous normal, No (%) | 5 (14.3) | 8 (36.4) |  |
| Burst-suppression, No (%) | 13 (37.1) | 4 (18.2) |  |
| Low voltage, No (%) | 2 (5.7) | 3 (13.6) |  |
| Flat trace, No (%) | 3 (8.6) | 2 (12.5) |  |
| Missing, No (%) | 4 (11.4) | 4 (18.2) |  |
| Neonatal seizures, No (%) | 29 (82.9) | 18 (81.8) | 1.00 |
| EEG confirmed, No (%) | 10 (54.3) | 8 (36.4) | 0.55 |
| Suspected (clinical), No (%) | 19 (54.3) | 10 (45.5) |  |
| Any epileptic drug received, No (%) | 32 (91.4) | 20 (90.9) | 1.00 |
| Phenobarbitone, No (%) | 32 (91.4) | 20 (90.9) | 0.24 |
| Midazolam, No (%) | 12 (34.3) | 9 (40.9) |  |
| Lidocaine, No (%) | 1 (2.9) | 4 (18.2) |  |
| Structural brain abnormality on MRI  at age 10-12y, No (%) | 24 (69%) | 3 (60%)^a^ | 1.00 |
| Basal ganglia/Thalamus, No (%) | 1 (3%) | 0 (0%)^a^ | NA |
| Watershed, No (%) | 14 (40%) | 2 (40%)^a^ | 1.00 |
| Solitary white matter lesions, No (%) | 8 (23%) | 0 (0%)^a^ | NA |
| Other, No (%) | 1 (3%) | 1 (20%)^a^ | 0.24 |
| WISC IV, full scale IQ at 6 to 8 years, median (IQR) | 102.5 (97 - 109) | 107.5 (104 - 111)^b^ | 0.35 |
| Any Neurologic/Neurodevelopmental  diagnose and/or IQ < 85, No (%) | 14 (40) | 7 (31.8)^c^ | 0.61 |

^a^Available for 5 (23%) of the children

^b^Available for 12 (55%) of the children

^c^At age 6-8 years

**Supplemental Figure 1** Flowchart of children included in the study with Sarnat grading of HIE severity in children exposed to Hypothermia-treated Hypoxic-Ischemic Encephalopathy (HIE).

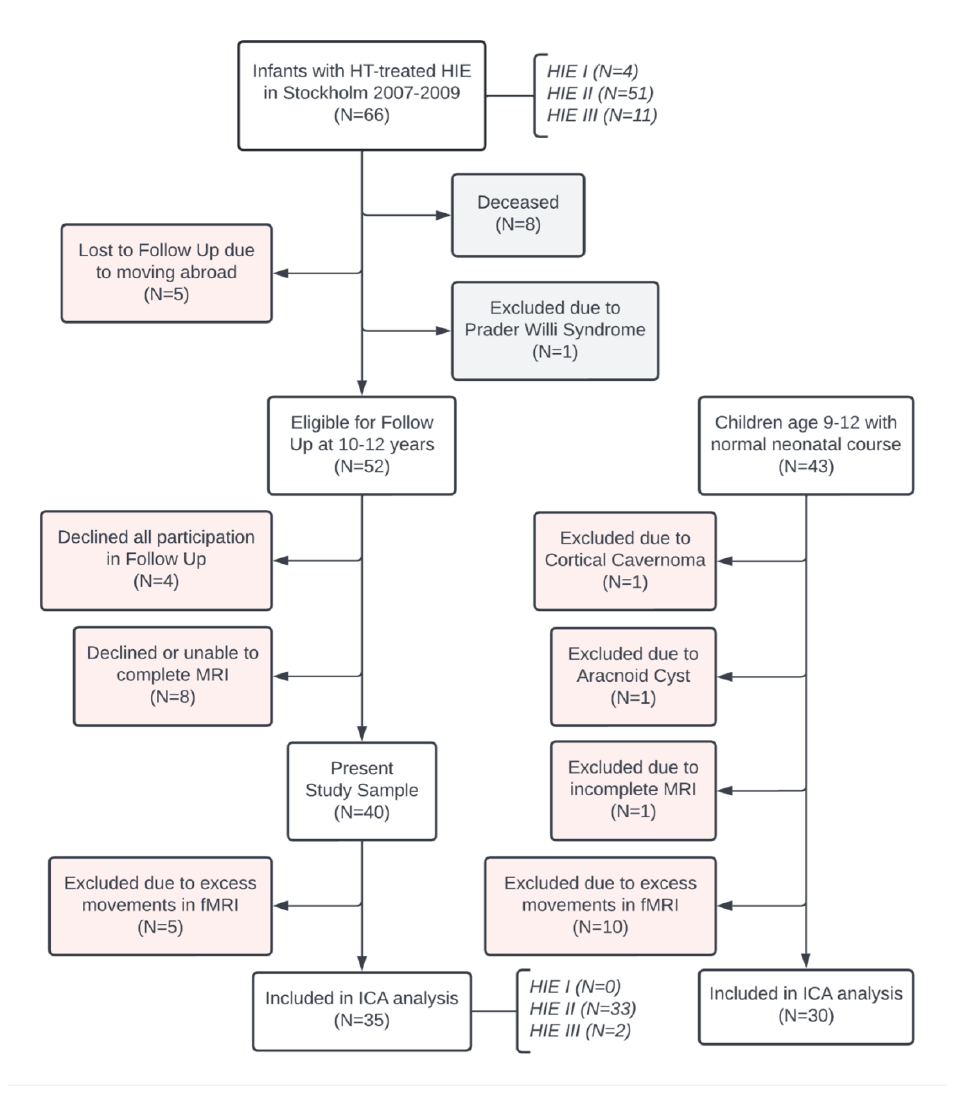

**Supplemental Figure 2** All 15 resting-state functional connectivity networks from a canonical ICA analysis with the data from the HIE and control subjects combined into a single dataset (fixed dimensionality of 20 components).

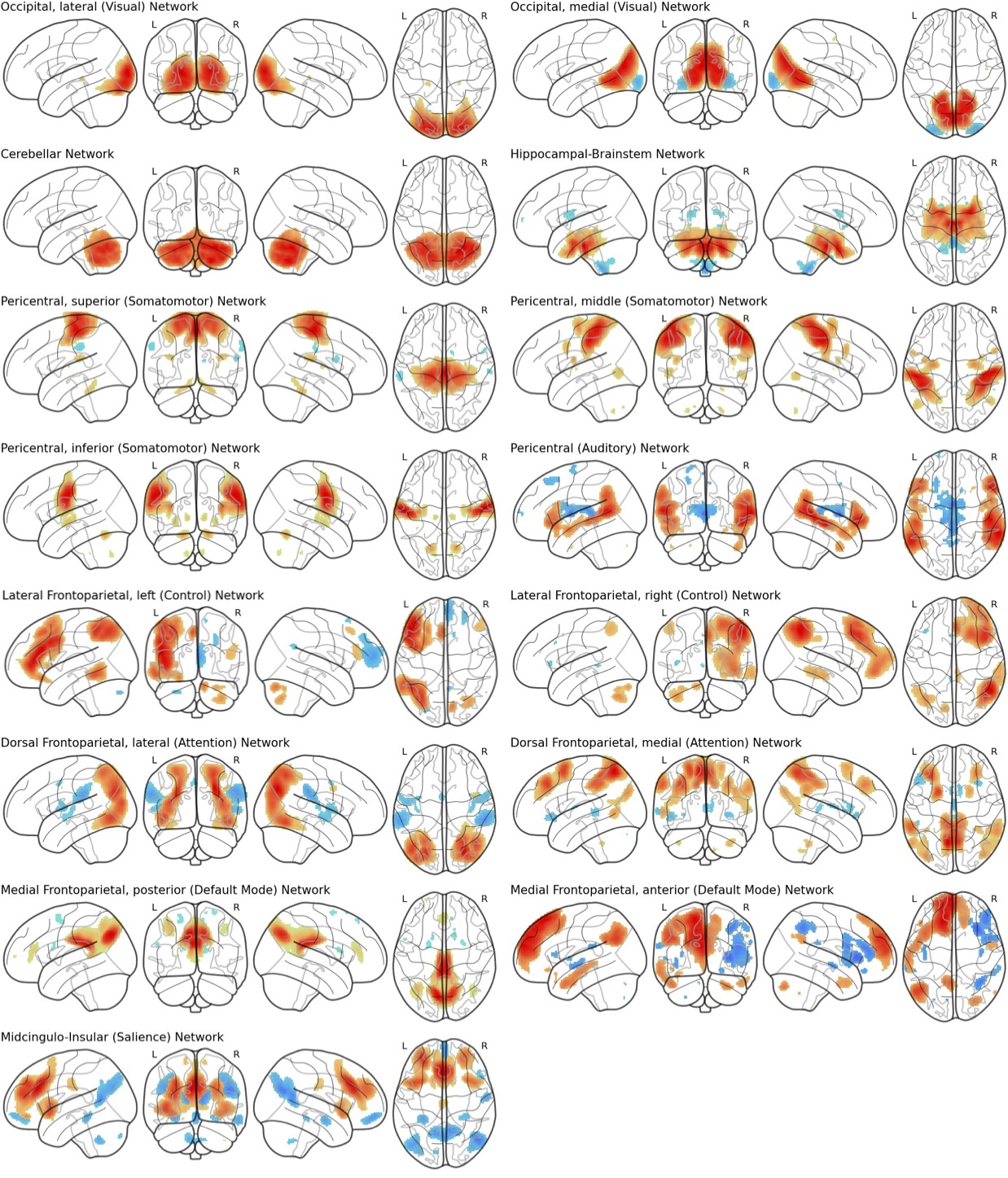

**Supplemental Figure 3. Differences in functional connectivity between groups with and without exposure for hypothermia-treated neonatal hypoxic-ischemic encephalopathy before correcting for age.** Results from non-parametric testing (p<0.05, voxel-wise threshold free cluster enhancement corrected (TFCE), minimal cluster size = 10) from data analysis with whole brain dual regression of the 15 ICA-derived networks (see Supplemental Figure 2) without covariates in the model detected clusters of decreased FC in children with hypothermia-treated HIE (N=35) compared to the control group (N=30) in four networks plotted on glass brain templates in A, with one cluster in the dorsolateral prefrontal cortex with decreased FC to the medial visual network plotted on anatomical template in B surviving statistical correction for multiple comparisons across all networks (Bonferroni, 15 two-sided tests, p<0.00167). All significant changes are colored red, seed networks are colored blue in A, MNI-coordinates are given in B.

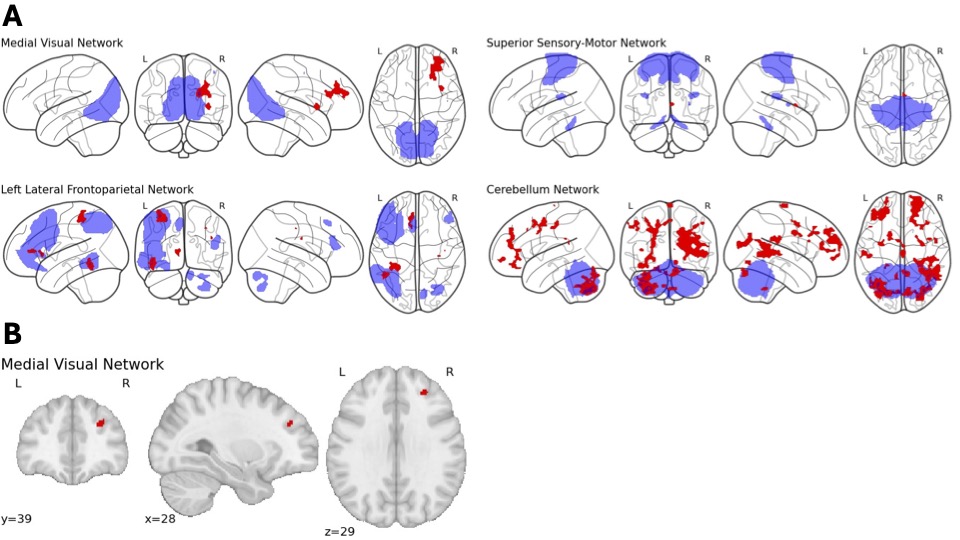
